# The *International Classification of Diseases, 10^th^ Edition, Clinical Modification (ICD-10-CM)* Code I16.0 Accurately Identifies Patients with Hypertensive Urgency

**DOI:** 10.1101/2023.02.05.23285422

**Authors:** Jed Kaiser, Vanessa Liao, Hooman Kamel, Catherine Ng, Richard I. Lappin, Ava L. Liberman

## Abstract

**Objective:** Hypertensive urgency, defined as acutely elevated BP without target organ damage, is associated with an increased risk of adverse cardiovascular events and accounts for a substantial proportion of national emergency department (ED) visits. To advance research in this space, we sought to validate the new *ICD-10-CM* diagnostic code for hypertensive urgency within a single healthcare system.

**Methods:** We performed a retrospective chart-review study of ED encounters at Weill Cornell Medicine from 2016 – 2021. We randomly selected 25 encounters with the *ICD-10-CM* code I16.0 as the primary discharge diagnosis and 25 encounters with primary *ICD-10-CM* discharge diagnosis codes for benign headache disorders. A single board-certified vascular neurologist reviewed all 50 encounters while blinded to the assigned *ICD-10-CM* codes to identify cases of hypertensive urgency. We calculated the sensitivity, specificity, and positive predictive values of the *ICD-10-CM* code I16.0 with 95% confidence intervals (CI).

**Results:** Out of 50 randomly selected ED encounters, 24 were adjudicated as hypertensive urgency. All encounters adjudicated as hypertensive urgency had been assigned the *ICD-10-CM* discharge diagnosis code of I16.0. All 25 of the encounters adjudicated as headache were assigned an *ICD-10-CM* discharge diagnosis code for a benign headache disorder. The *ICD-10-CM* code for hypertensive urgency, I16.0, was thus found to have a sensitivity of 100% (95% CI: 86-100%), specificity of 96% (95% CI: 80-100%), and positive predictive value of 96% (95% CI: 78-99%).

**Conclusion:** We found that the new *ICD-10-CM* code for hypertensive urgency, I16.0, can reliably identify patients with this condition.

## Introduction

Uncontrolled hypertension is a leading cause of cardiovascular disease and premature death worldwide.^1^ The prevalence of hypertension in the US is substantial with nearly half of all adults (47% or 116 million people) diagnosed with hypertension.^2^ Recent studies have noted that blood pressure (BP) control among hypertensive adults has decreased in recent years.^3^ Unsurprisingly, the national proportion of emergency department (ED) visits for hypertensive crises has increased steadily.^4,5^ An estimated 40.4 million patients seen between 2016-2019 in US EDs had severe hypertension, equating to 6.1% (95% CI: 5.7-6.5%) of all ED visits.^6^

Health services researchers interested in acutely hypertensive patients have mainly focused on instances where hypertension results in target-organ damage rather than hypertensive urgency, defined as acutely elevated BP without immediately apparent target organ damage, despite the fact that the latter is associated with substantial future cardiovascular risk.^7^ One reason for this research discrepancy may be that, unlike hypertensive patients with target-organ damage,^8^ identifying hypertensive urgency patients using administrative claims data has been challenging as, until recently, there were no diagnostic codes for this condition. With the adoption of the *International Classification of Diseases and Related Health Problems, 10*^*th*^ *Edition-Clinical Modifications (ICD-10-CM)* in the US in 2015, a diagnostic code specifically for hypertensive urgency, I16.0, was introduced. To advance acute hypertension research we herein seek to determine the validity of the new *ICD-10-CM* for hypertensive urgency; we are not aware of any prior studies evaluating this diagnostic code.

## Methods

### Design and Setting

We conducted a retrospective study of ED encounters at the two sites of a single hospital center, Weill Cornell Medicine (WCM), from 1/1/2016 to 12/1/2021. One study site is a community-based 27-bed adult ED whereas the other is a 50-bed ED that is part of a tertiary referral center. Both study sites are located in New York City, New York.

### Study Population

To validate the new *ICD-10-CM* code for hypertensive urgency, we identified ED encounters with the code I16.0 as the primary discharge diagnosis. As a comparator, we also identified encounters with a primary *ICD-10-CM* discharge diagnosis code for a benign headache disorder: G43-G43.5x, G43.7x-G43.9x, G43.A-G43.D, G44, or R51. These diagnostic codes for headache have been previously used in administrative claims research.^9^ We selected patients with a primary headache disorder as a comparator since headache complaints are very common in patients with severely elevated blood pressure^6^ and patients with headache complaints often have elevated blood pressure.^10^ For this study, we only included ED encounters that resulted in discharge to home (ED treat-and-release visit). To identify relevant encounters, we leveraged our existing institutional infrastructure for secondary use of data from the existing electronic health record (EHR).^11^

As in a prior *ICD* code validation study,^12^ we randomly selected 25 encounters with a primary discharge diagnosis of I16.0 and 25 encounters with a primary discharge diagnosis of any of the aforementioned *ICD-10-CM* benign headache disorder codes. A single board-certified vascular neurologist (A.L.L.) reviewed detailed medical records of all 50 selected encounters while fully blinded to their assigned *ICD-10-CM* discharge diagnosis codes. Determination of hypertensive urgency was based on its standard definition as severely elevated BP without evidence of target organ damage; no specific BP measurement criteria were used during the adjudication process given the variable definition of severely elevated BP in extant literature as well as clinical practice.^13,14^ Institutional review board approval was obtained for this study from the WCM Institutional Review Board which granted a waiver of informed consent in accordance with the Declaration of Helsinki (reference number = 22-12-071-380).

### Statistical Analysis

We used standard descriptive statistics, including medians with interquartile ranges (IQR), to describe all included encounters, abstracting pre-specified demographic and clinical variables from the EHR. We compared patients with an adjudicated diagnosis of headache versus hypertensive urgency using two-sided chi-squared test for dichotomized variables and Mann-Whitney U for continuous variables. Using the adjudicated diagnosis based on detailed medical record review as the gold standard, we calculated the sensitivity, specificity, and positive predictive value with 95% confidence intervals (CI) of *ICD-10-CM* code I16.0. We considered P < 0.05 to be statistically significant. Analyses were performed using Stata/MP, version 15.1 (StataCorp, TX).

## Results

Of the 50 selected ED encounters, blinded adjudication using the EHR resulted in a clinical diagnosis of hypertensive urgency for 24 encounters and a benign headache disorder for 25 encounters. One encounter was adjudicated as a transient ischemic attack (TIA) and was included in the non-hypertensive urgency category for our analyses. The median age of the entire cohort was 59 years (IQR: 50-69) and 62% were female.

Comparing patients with hypertensive urgency (n=24) to those with a headache disorder (n=25), those with hypertensive urgency were older (64 vs. 54 years of age; P = .04), more likely to have a history of hypertension (83% vs. 28%; P < .001), and less likely to have a history of a headache disorder prior to ED arrival (4% vs. 40%; P = .002). Sex and race were similar between the two groups (Table 1).

**Table 1:**
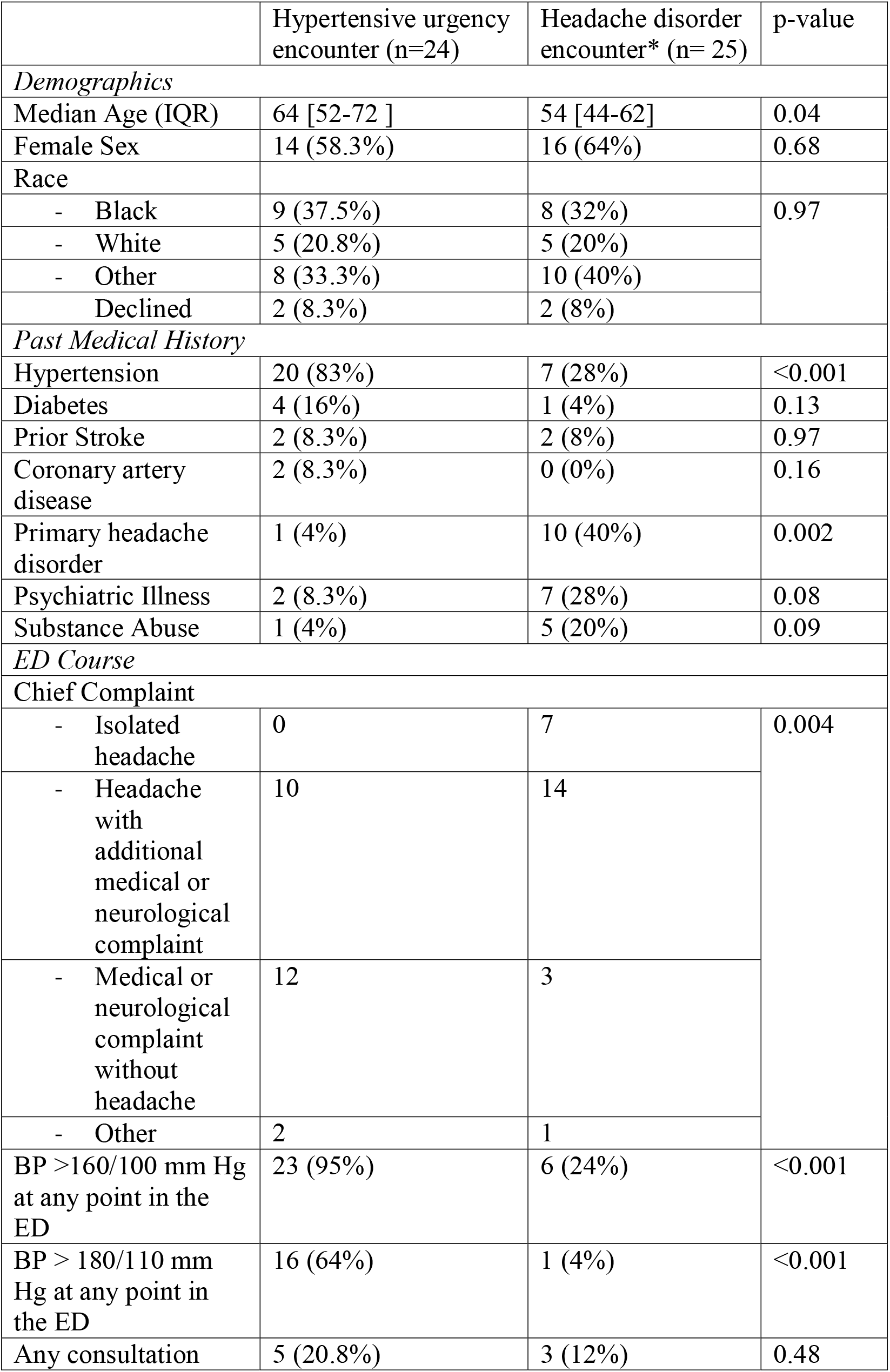
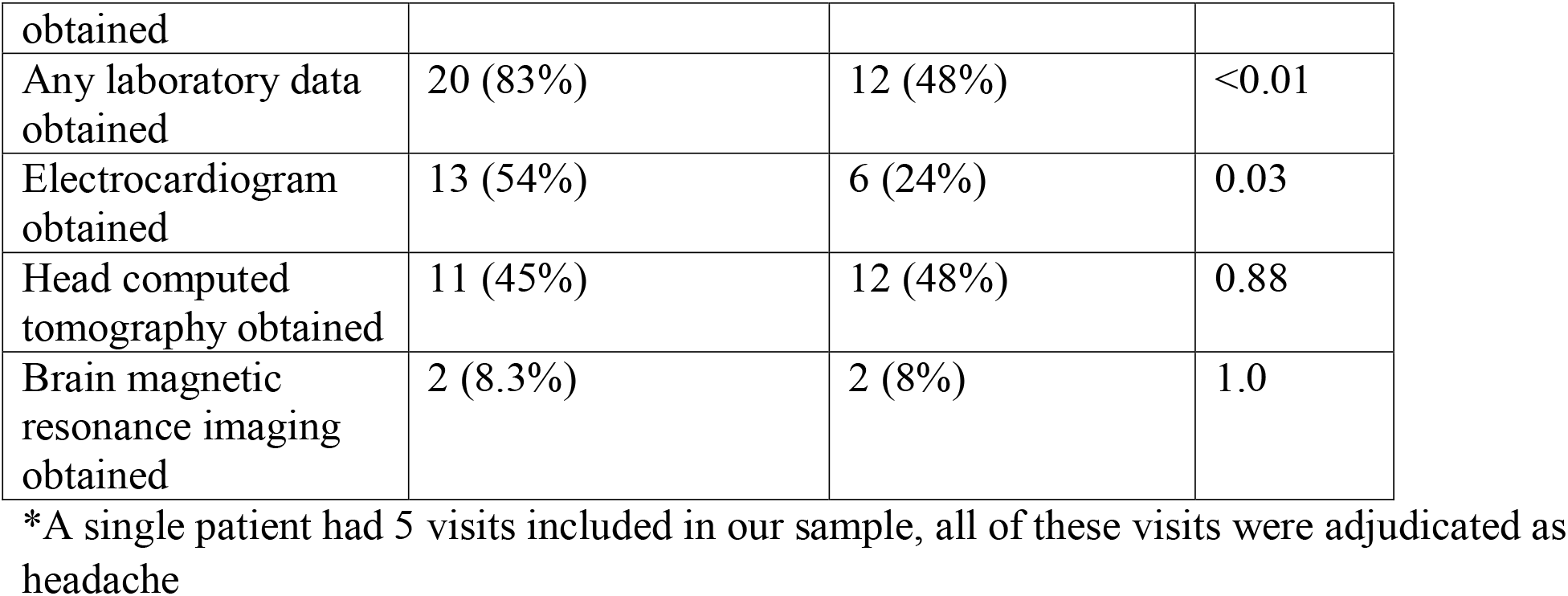
Included encounters adjudicated as having an ED treat- and-release visit for headache or hypertensive urgency (N= 49)

A chief complaint of isolated headache was exclusively seen in patients with a primary headache disorder whereas patients with hypertensive urgency often had neurological and/or medical complaints that did not include headache. Among patients with hypertensive urgency, 95% had a measured BP >160/100 mm Hg and 64% had a BP >180/110 mm Hg during their ED visit. Patients with hypertensive urgency were more likely to have laboratory testing (83% vs. 48%; P < .01) as well as an electrocardiogram (54% vs. 24%; P = .03) obtained in the ED than headache patients. Head computed tomography and brain magnetic resonance imaging were obtained with similar frequency among included headache and hypertensive urgency patients (Table 1).

All 24 of the encounters adjudicated as hypertensive urgency had been assigned an *ICD-10-CM* discharge diagnosis of I16.0 and all 25 of the encounters adjudicated as headache were assigned an *ICD-10-CM* discharge diagnosis code for a benign headache disorder. The one encounter that was determined to be a TIA was assigned an *ICD-10-CM* code of I16.0. Thus, the *ICD-10-CM* code for hypertensive urgency, I16.0, had a sensitivity of 100% (95% CI: 86-100%), a specificity of 96% (95% CI: 80-100%), and positive predictive value of 96% (95% CI: 78-99%; Table 2).

**Table 2.**
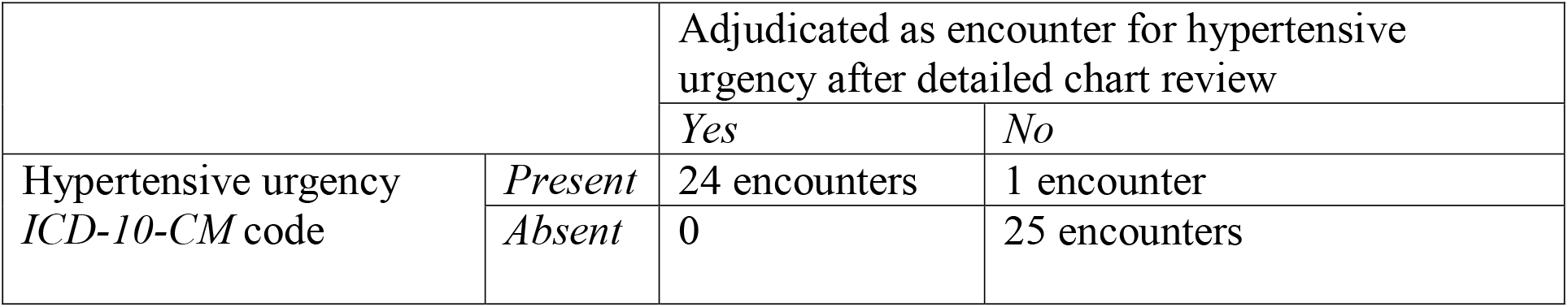
Hypertensive Urgency *ICD-10-CM* Code Validation

## Discussion

In this retrospective cohort study, the new *ICD-10-CM* code for hypertensive urgency, I16.0, was found to be highly accurate. Based on our findings, this code can be used to identify patients with hypertensive urgency, particularly in the emergency setting, in future research using administrative claims data. The ability of the *ICD-10-CM* code for hypertensive urgency to identify patients with this condition is similar to that of other *ICD* codes for cardiovascular diseases.^12,15^ For example, the *ICD* code for accurately identifying ED patients who suffered an out-of-hospital cardiac arrest was shown to have a specificity of 99%, sensitivity of 87%, and positive predictive value of 92%.^15^

The primary strength of this study is its use of blinded, expert adjudication to determine the validity of the new *ICD-10-CM* code as compared to patients with similar clinical characteristics. Important limitations include the fact that, since variable BP measurements are reported in the literature to define hypertensive urgency,^13,14^ we relied on provider notes to adjudicate the diagnosis rather than BP measurements alone. However, our broad definition of hypertensive urgency as severely elevated blood pressure without target-organ damage is consistent with clinical practice and the majority of patients had BP >160/100 mm Hg. Additionally, since we only included encounters where patients were discharged to home from the ED in this study, so as not to bias case adjudication, additional research to determine the validity of the new hypertensive urgency code in the inpatient or outpatient setting may be warranted. Lastly, our results are from a single hospital center which limits the generalizability of our findings.

## Conclusion

We found that the *ICD-10-CM* code for hypertensive urgency, I16.0, had a sensitivity of 100%, specificity of 96%, and positive predictive value of 96% as compared to detailed chart review.

## Data Availability

All data generated or analyzed during this study are included in this published article.

## DECLARATIONS

### Ethics Approval and Consent to Participate

Institutional review board approval was obtained for this study from the WCM Institutional Review Board which granted a waiver of informed consent.

### Consent for Publication

All authors have read and approved the submitted manuscript for publication.

### Availability of Data and Materials

All data generated or analyzed during this study are included in this published article.

